# Prediction Models for Severe Manifestations and Mortality due to COVID-19: A Rapid Systematic Review

**DOI:** 10.1101/2021.01.28.21250718

**Authors:** Jamie L. Miller, Masafumi Tada, Michihiko Goto, Nicholas Mohr, Sangil Lee

## Abstract

**Background:** Throughout 2020, the coronavirus disease 2019 (COVID-19) has become a threat to public health on national and global level. There has been an immediate need for research to understand the clinical signs and symptoms of COVID-19 that can help predict deterioration including mechanical ventilation, organ support, and death. Studies thus far have addressed the epidemiology of the disease, common presentations, and susceptibility to acquisition and transmission of the virus; however, an accurate prognostic model for severe manifestations of COVID-19 is still needed because of the limited healthcare resources available.

**Objective:** This systematic review aims to evaluate published reports of prediction models for severe illnesses caused COVID-19.

**Methods:** Searches were developed by the primary author and a medical librarian using an iterative process of gathering and evaluating terms. Comprehensive strategies, including both index and keyword methods, were devised for PubMed and EMBASE. The data of confirmed COVID-19 patients from randomized control studies, cohort studies, and case-control studies published between January 2020 and July 2020 were retrieved. Studies were independently assessed for risk of bias and applicability using the Prediction Model Risk Of Bias Assessment Tool (PROBAST). We collected study type, setting, sample size, type of validation, and outcome including intubation, ventilation, any other type of organ support, or death. The combination of the prediction model, scoring system, performance of predictive models, and geographic locations were summarized.

**Results:** A primary review found 292 articles relevant based on title and abstract. After further review, 246 were excluded based on the defined inclusion and exclusion criteria. Forty-six articles were included in the qualitative analysis. Inter observer agreement on inclusion was 0.86 (95% confidence interval: 0.79 - 0.93). When the PROBAST tool was applied, 44 of the 46 articles were identified to have high or unclear risk of bias, or high or unclear concern for applicability. Two studied reported prediction models, 4C Mortality Score from hospital data and QCOVID from general public data from UK, and were rated as low risk of bias and low concerns for applicability.

**Conclusion:** Several prognostic models are reported in the literature, but many of them had concerning risks of biases and applicability. For most of the studies, caution is needed before use, as many of them will require external validation before dissemination. However, two articles were found to have low risk of bias and low applicability can be useful tools.

## INTRODUCTION

### Background

COVID-19 is the disease caused by SARS-CoV-2 that emerged in China in December 2019.^1^ Throughout 2020, COVID-19 has become an increasing threat to public health on national and global level.^2^ There has been an immediate need for research to help predict the clinical deterioration including death, mechanical ventilation, and organ support.^3^ A precise risk stratification of individuals with COVID-19 enables allocation of appropriate resources such as hospitalization, intensive care admission, ventilatory support, and antiviral therapy.^4^ As we start to understand the unique nature of this infection, an accurate risk stratification tool is urgently needed.

Studies thus far have addressed the epidemiology of the disease, common presentations, and susceptibility to disease acquisition and transmission of the virus.^5,6,7^ However, the short- and long-term implications of developing severe illnesses caused by COVID-19 is still widely unknown. Many studies with methodological shortcomings are published to facilitate the dissemination of rapidly evolving science of COVID-19.^8,9^ The quality of prediction models related to COVID-19 also suffered, as many studies did not have adequate validation steps after models were developed.^9^ The purpose of this review is to consolidate published data on validated predictive models of severe illnesses caused by COVID-19. We undertook a rapid systematic review following the format given by the King et al^10^

### Goals of this investigation

The goal of our review was to collectively evaluate published findings of prediction models for severe illness caused by COVID-19 to inform healthcare providers and the general population of the existing evidence in the midst of the pandemic.

## METHODS

The review protocol was registered to PROSPERO (registration number CRD42020201484). Our study adheres to the Preferred Reporting Items for Systematic Reviews and Meta-analyses guidelines for systematic reviews and was performed in accordance with best-practice guidelines (PRISMA).^11^

Search strategies were developed with the assistance of a health sciences librarian with expertise in systematic reviews. Searches were developed by the primary author and librarian using an iterative process of gathering and evaluating terms. Searches were finalized in July 2020. Comprehensive strategies, including both index and keyword methods, were devised for PubMed and EMBASE. In order to maximize sensitivity, no pre-established database filters were used other than English language. We also screened references of articles identified by these systematic search strategies to look for any additional records potentially useful for the aim of this review. Lastly, we examined additional articles through a referral from investigators and online journal updates. The completed PubMed strategy is shown in Supplemental file 1. PubMed Strategy was then adapted for EMBASE and is available upon request. The last search was conducted on 7/27/2020.

We included studies of randomized controlled trials, cohort studies, and case-control studies that discuss the possible short-term and long-term consequences of contracting COVID-19 in any clinical setting. All studies were considered regardless of publication status (preprint or peer review manuscript), as long as those are included to PubMed or EMBASE. The definition of positive COVID-19 included a positive nasopharyngeal or oropharyngeal swab for COVID-19 by polymerase-chain reaction (PCR) or antigen tests, serological test for COVID-19, or those who were clinically diagnosed as COVID-19 based on clinical presentation and epidemiologic information. Outcomes included intubation, mechanical ventilation, any other type of organ support (for example, hemodialysis), and/or death. We included studies that developed and validated a multivariable model or scoring system, based on individual participant level data, to predict any COVID-19 related outcome or externally validated a known scoring system. We excluded review articles, case reports, editorials, and comments. Studies reporting the predictive ability of a single test, for example, a study only focused on the prognostic value of lactate dehydrogenase (LDH), were excluded from the review, since we were primarily interested in the combination of test results with any internal or external validation. Epidemiological studies that aimed to model disease transmission or diagnostic test accuracy, and predictor finding studies were also excluded.

### Data Collection and Processing

Titles, abstracts, and full texts were screened in duplicate for eligibility by independent reviewers (JM and SL), and discrepancies were resolved through discussion moderated by the third reviewer (MG). The reasons for any exclusions were recorded for those that required full article review.

Two investigators (JM, MT) independently extracted data from the included studies. The investigators underwent initial training and extracted data into a predesigned data collection form. The following information was abstracted: author, year, setting, outcome, predictors in final model, sample size for development set, type of validation, sample size, performance, and geographical region.

When data were missing or ambiguous, we contacted the authors for clarification. Studies were independently assessed for risk of bias by two investigators (JM, MT) using the Prediction Model Risk Of Bias Assessment Tool (PROBAST) tool. Any discrepancies were resolved by consensus.

### Primary Data Analysis

A narrative synthesis of the characteristics of the included studies, including the outcomes, sample size for development and validation, type of case ascertainment, relationship between the final model, and the development of severe illnesses, were summarized in a table. We listed risk of bias and applicability using the PROBAST^12^ tool. To maximize the synthesis of results, we reported the studies based on the low risk of bias, low concern for applicability, and geographical location for future use.

## RESULTS

### Study selection

We included a total of 292 of 3399 articles based on the title and abstract. Then, we included a total of 42 out of 292 during the secondary full text screening up to July 2020. We identified an additional four articles from an investigator, online journal update, and references. The details of the selection process are listed in Figure 1. Overall, kappa was 0.86 (95% 0.79 - 0.93) between two reviewers.

**Figure 1.**
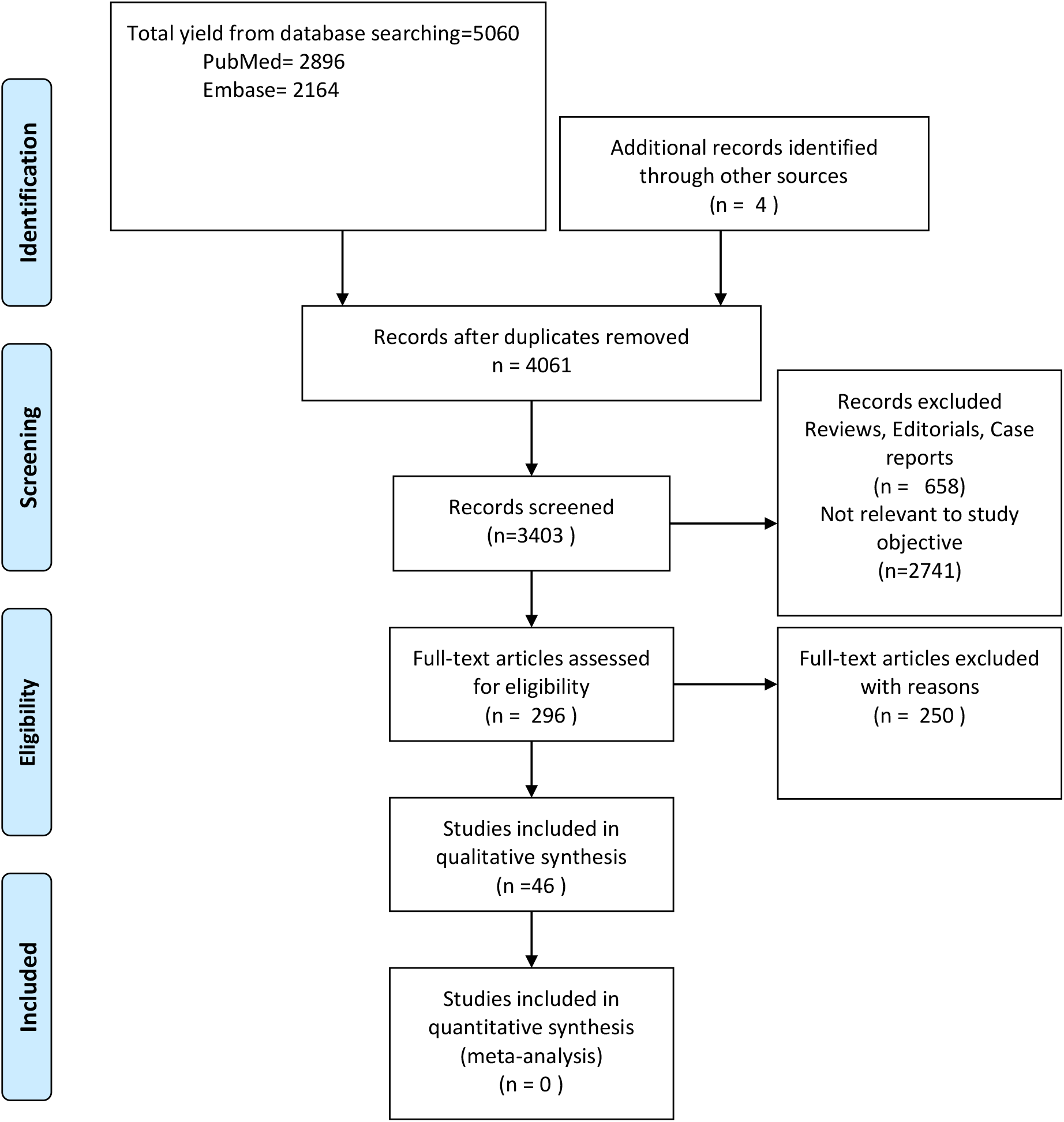
PRISMA diagram.

### Characteristics of studies

A total of 41 (89%) articles were identified from peer review journals and 5 (11%) were from preprints (Figure 1). The majority of studies (n=25, 53%) were published in China, 9 studies (19%) in the U.S. and 7 (15%) were published in Europe. (Table 1).

**Table 1.** Data Summary.

| First Author | Type of Study | Outcome | Prediction Model | Sample Size | Type of validation | Performance (AUC) | Location |
| --- | --- | --- | --- | --- | --- | --- | --- |
| Al-Najjar H et al.<sup>18</sup> | Retrospective | Death | Country, infection reason, sex, group, confirmation date, birth year, and region | 659 | Split | .908 for recovery, .985 for death | Korea |
| Bae J et al.<sup>19</sup> | Retrospective | Mechanical ventilation, death | Chest X-Ray (deep learning) | 514 | Split | .904 for ventilation<br>.936 for mortality | US |
| Bahl et al.<sup>20</sup> | Retrospective | Death | Age, RR, SpO2, Cr, ALT, procalcitonin, lactic acid | 1629 | LOOCV | V 0.80 | US |
| Bello-Chavolla, O. Y. et al.<sup>21</sup> | Retrospective | Death | Age, diabetes, diabetes, obesity, CKD, hypertension, and immunosuppression | 177 133 | Split | V .823 | Mexico |
| Bi X et al.<sup>22</sup> | Retrospective | Severe case | Fibrinogen to albumin ratio, platelet | 141 | External validation | V .754 | China |
| Borghesi A et al.<sup>23</sup> | Retrospective | Death | Brixia score, age, immunosuppression | 302 | External validation | V .853 | Italy |
| Chen R et al.<sup>24</sup> | Retrospective | Death | Age, CHD, CVD, dyspnea, procalcitonin, AST | 1590 | Bootstrap | V .91 (95% CI, .85-.97) | China |
| Chen A et al.<sup>25</sup> | Retrospective | Death | BUN, D-dimer | 305 | Bootstrap | V 0.929 | China |
| Cheng F et al.<sup>26</sup> | Retrospective | ICU | RR, WBC, Lymphocyte, DBP, CRP, SpO2, age, temp, pulse, QRS duration, BUN, sodium, T wave axis, anion gap, SBP, PR interval, R wave axis, RBC, Calcium, Albumin | 1987 | Split | V .799 (95% CI: .752-.846) | US |
| Clift A et al.<sup>14</sup> | Retrospective | Death | age, ethnicity, deprivation, BMI, comorbidity | 8256158 | Two temporary validations | V 0.928 (0.919 to 0.938) | UK |
| Dong Y et al.<sup>27</sup> | Retrospective | Death | HTN, neutrophil to lymphocyte, and NT-Pro BNP | 628 | Split | V 0.922 (14day), V 0.881 (21 day) | China |
| Galloway J et al.<sup>28</sup> | Retrospective | ICU, death | age, male, non-white ethnicity, SpO2, radiological severity score>3, neutrophil count, CRP, albumin, Cr, diabetes, hypertension and chronic lung disease | 1157 | Split | V .714 (95%CI .665, .762) | UK |
| Gong J et al.<sup>29</sup> | Retrospective | Severe case | Age; LDH, CRP, RDW, BUN, and direct bilirubin; and albumin | 372 | Split | V .853 (95% CI, .790-.916) | China |
| Haimovich et al.<sup>30</sup> | Retrospective | Respiratory failure | Quick COVID severity index (RR, SpO2, and O2 flow rate), COVID severity score (O2 flow, SpO2, AST, Chloride, Procalcitonin, RR, SBP, ALT, WBC, BUN, Glucose, CRP, Ferritin, age, Cr) | 1792 | 10-fold cross validation | V.81 (95% CI.73, .89) for the quick COVID severity index, V 0.76 [95% CI 0.65 to 0.86] for COVID severity score | US |
| Hu H et al.<sup>31</sup> | Retrospective | Death | Rapid Emergency Medicine Score (REMS) | 138 | External | V 0.833 (95% confidence interval [CI] = 0.737 to 0.928) | China |
| Jang JG et al.<sup>32</sup> | Retrospective | Death and critical outcomes | SIRS, qSOFA, NEWS | 110 | External | V SIRS = .639 (95% CI, .423-.856) qSOFA = .779 (95% CI, .600-.957) NEWS = 0.867 (95% CI .709-1.000) for mortality, SIRS = .744 (95% CI, .602-.886), qSOFA = .760 (95% CI, .620-.899), and NEWS = .918 (95% | Korea |
|  |  |  |  |  |  | CI, .841–.995) for critical outcomes |  |
| Ji D et al.<sup>33</sup> | Retrospective | Severe case | Age, comorbidity, lymphocyte, LDH | 208 | bootstrap | V .91 (95% CI, .86-.94) | China |
| Knight et al.<sup>12</sup> | Retrospective | Death | age, sex, comorbidities, RR, SpO2, level of consciousness, BUN, and CRP | 57824 | Split | V range .767 (95%CI .760, .773) | UK, Wales, Ireland |
| Li M et al.<sup>34</sup> | Retrospective | Mechanical ventilation, death | Pulmonary X ray severity score | 263 | External | V .80 (95%CI .75-.85) | US |
| Li Q et al.<sup>35</sup> | Retrospective and prospective | Severe disease | Age, LDH, CD4 | 639 | Prospective | V .92 | China |
| Liang W et al.<sup>36</sup> | Retrospective | Mechanical ventilation, death, ICU | chest radiographic abnormality, age, hemoptysis, dyspnea, unconsciousness, number of comorbidities, cancer history, neutrophil-to-lymphocyte ratio, LDH and direct bilirubin. | 2300 | Split | D .88 (95% CI, .85-.91)<br>V .88 (95% CI, .84-.93) | China |
| Liu Y et al.<sup>37</sup> | Retrospective | Severe/Critical illness | Neutrophil to lymphocyte ratio, CRP | 84 | Split | D .804 (95% CI: .702-.883),<br>V .881 (95% CI: .782-.946) | China |
| Matos J et al.<sup>38</sup> | Prospective | Mechanical ventilation, death | Gender, chronic lung disease, duration of symptom, predominant type, WBC count, aortic and coronary calcification, comorbidity, lymphocyte, age, volume of disease, CRP | 106 | Cross validation | V 0.92 | Italy |
| McRae et al.<sup>39</sup> | Prospective | Death | biomarkers (cTnI, PCT, MYO, and CRP), age, and sex | 172 | External validation | V 0.94 (0.89–0.99) | China |
| Myrstad et al.<sup>40</sup> | Prospective | Severe disease | NEWS2 score≥6 | 66 | External validation | V .822, (95% CI .690-.953) | Norway |
| Laguna-Goya et al.<sup>41</sup> | Prospective | Death | SpO<sub>2</sub>/FiO<sub>2</sub> ratio, neutrophil-to-lymphocyte ratio, LDH level, IL-6 level, and age | 611 | bootstrap | V.93 | Spain |
| Satici C et al.<sup>42</sup> | Retrospective | Death | PSI, CRP | 681 | External validation | V .92 (95% CI .89-.94) | Turkey |
| Shang Y et al.<sup>43</sup> | Retrospective | Death | Age, CVD, lymphocytes, Procalcitonin and D-dimer | 452 | External validation | V .938(95%CI .902-.973) | China |
| Shashikumar SP et al.<sup>*44</sup> | Retrospective and prospective | Intubation | Deep learning<sup>#</sup> | 777 | External validation | V.918 | US |
| Singh K et al.<sup>*45</sup> | Retrospective | Death, ICU, intubation | Epic deterioration index | 174 | External validation | V0.67 (95% CI 0.59-0.75) | US |
| Sun H et al.<sup>*46</sup> | Retrospective and prospective | Hospitalization, critical illness, death | CoVAS model<sup>§</sup> | 11,586 | Cross validation and External | Hospitalization V.76, critical illness V.79, death V.93 | US |
| Sun L et al.<sup>47</sup> | Retrospective | Severe and critical symptoms | age, GSH, CD3 percentage and total protein | 336 | Not specified | D 0.9997 and V 0.9757 | China |
| Wang B et al.<sup>48</sup> | Retrospective | Death | Age, Chest tightness, AST, BUN | 104 | Bootstrap | V.893 (95% CI, .807-.980) | US |
| Wang K et al.<sup>49</sup> | Retrospective | Death | Age, HTN, CAD (clinical)<br>Age, CRP, SpO<sub>2</sub>, Neutrophil, Lymphocyte, d-dimer, AST, GFR (lab based) | 340 | Split | Clinical V .83 (95%CI .68-.93),<br>Lab V .88 (95%CI .75-.96) | China |
| Wei W et al.<sup>50</sup> | Retrospective | Severe COVID-19 | CT score, CD8, CRP, tight chest | 81 | LGOCV | Clinical D.97 V.90<br>Radiomics D.95 V.89 | China |
| Wollenstein-Betech S* et al.<sup>15</sup> | Retrospective | 1)Hospitalization, 2)death, 3)ICU, 4)mechanical ventilation | 1)age, gender, CKD, diabetes, immunosuppression, 2) age, SARS-CoV-2 test, immunosuppression and pregnancy, 3)pneumonia, CVD, asthma, and SARS-CoV-2 test, 4)ICU and pneumonia, age, gender, CVD, obesity, pregnancy, and SARS-CoV-2 test | 91,179 | Cross validation | 1).612-.622<br>2).674-.687<br>3).538-.554<br>4).541-.560 | Mexico |
| Wu G et al.<sup>13</sup> | Retrospective | Severe or critical illness | Age, Lymphocytes, CRP, LDH, CK, BUN, Ca, Total protein | 725 | Prospective validation | V range of .84 to .93 | China, Italy, Belgium |
| Wu Q et al.<sup>51</sup> | Retrospective | death, mechanical ventilation, ICU | clinic-radiomics signature (CrrScore) early/late phase | 492 | Split | Early phase<br>D .826 (.714-.937),<br>V .850 (.763-.935),<br>Late phase<br>D .911 (.796-.999),<br>V .886 (.675-.999) | China |
| Wu S et al.<sup>52</sup> | Retrospective | Severe COVID-19 | Age, neutrophil, lymphocyte, procalcitonin, and C-reactive protein | 270 | Split | D .955<br>V .945 | China |
| Xiao, S. et al.<sup>53</sup> | Retrospective | Severe case | Hypertension, Neutrophil, CRP, Lymphocyte, LDH | 690 | Internal and external validation | V .871, 95% CI: .769-.972 (Internal),<br>V .826, 95% CI: .746-.907 (external) | China |
| Yadaw A et al.*<sup>54</sup> | Retrospective and prospective | Death | Age, SpO2, type of encounter, hydroxychloroquine, and max temperature | 5051 | External | V .91 | US |
| Zhang C et al.<sup>3</sup> | Retrospective | Severe COVID-19 | Age, WBC, Neutrophil, GFR, Myoglobin | 80 | Scoring validated internally | V .906 | China |
| Zhang S et al.<sup>55</sup> | Retrospective | Death | Age, LDH, Neutrophil to lymphocyte, and bilirubin | 828 | External | V<br>#1 .878,<br>#2 .839 | China |
| Zheng Y et al.<sup>56</sup> | Retrospective | Composite outcome ICU, mechanical ventilation, death<br>Secondary Death | Age, sex, comorbidity, lymphocyte, extent, crazy paving sign, change in liver density | 238 | Split | V .89 (95% CI, .82–.96) | China |
| Zheng Y et al.<sup>57</sup> | Retrospective | Severe COVID-19 | Neutrophil, Lymphocytes, and Platelets (NLP score) | 141 | Validation with survival nomogram | V .821 (95% CI, .746–.896) | China |
| Zhou Y et al.<sup>58</sup> | Retrospective | Severe COVID-19 per ATS guidelines | temperature, cough, dyspnea, HTN, CVD, CLD, and CKD | 366 | bootstrapping | Derivation.86<br>Validation.84<br>Nomogram .86 | China |

\*Preprint only, # Deep learning model does not give an importance of variables for prediction. \$ Model variables include age, diastolic BP, SpO2, Positive COVID, respiratory rate, stroke, chest x ray finding, heart rate, temperature, systolic BP, Charlson score, intracranial hemorrhage, subarachnoid hemorrhage, male, hematologic malignancy, renal cancer, pancreatitis, cystic fibrosis, cardiac arrest, seizure, amyolateral sclerosis, metabolic acidosis, myasthenia gravis, pneumothorax, spinal muscular dystrophy, pericarditis, high or low BMI, ARDS. D: Derivation AUC, V: Validation AUC.

There were 38 (82%) retrospective cohort studies, and four (9%) prospective cohort studies, and four (9%) retrospective and prospective studies. The majority (n=25, 54%) were conducted at a single institution, 21 studies (46%) were conducted at two or more sites. There were two studies that were conducted in multiple nations.^12, 13^ The total sample was 8,621,479 patients, and the mean sample size used for model derivation in the 46 articles was 187,423 (range 66-8,256,158).

### Critical appraisal of quality

Next, using the PROBAST, we evaluated the risk of bias for each study and concern for applicability using the low, high, or unclear categories. The articles where the PROBAST tool did not apply were categorized as not applicable.

The PROBAST composite scores of the 46 articles, at the domain and item level, are presented in Figure 2. The overall risk of bias was low in four (9%) studies, high in 34 (74%) studies, and unclear in eight (17%) studies. The overall concern for applicability was low in 22 (48%) studies, high in 17 (37%) studies, and unclear in seven (15%) studies. (Figure 2) The lowest risk of bias was seen in the participants domain (29 studies with low risks of bias, 63%), and highest risk of bias was seen in the analysis domain (32 studies with high risks of bias, 70%). The lowest concern for applicability was seen in the participants domain (38 studies with low concern, 83%), and the highest risk of concern for applicability was seen in the outcome domain (high concern, n=16, 35%). When the PROBAST evaluation was stratified to multi-site studies, there were increase in the overall applicability (Figure 3).

**Figure 2.**
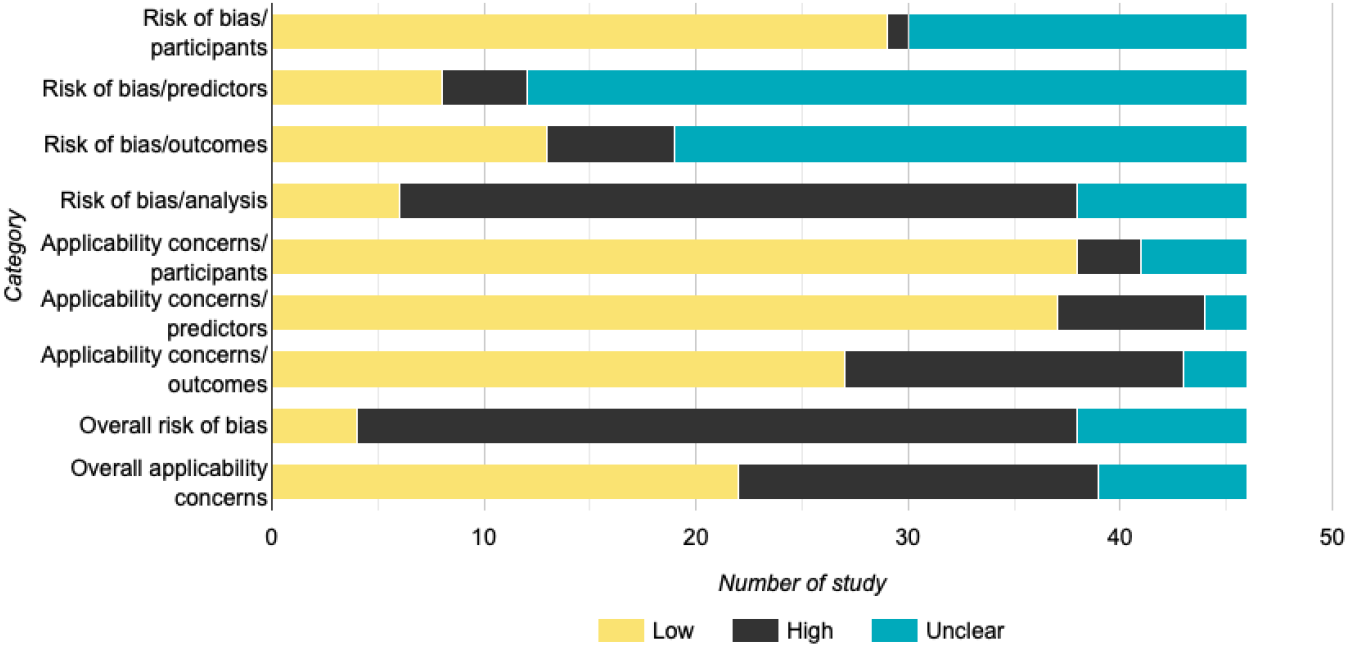
Risk of Bias and Concern for Applicability

**Figure 3.**
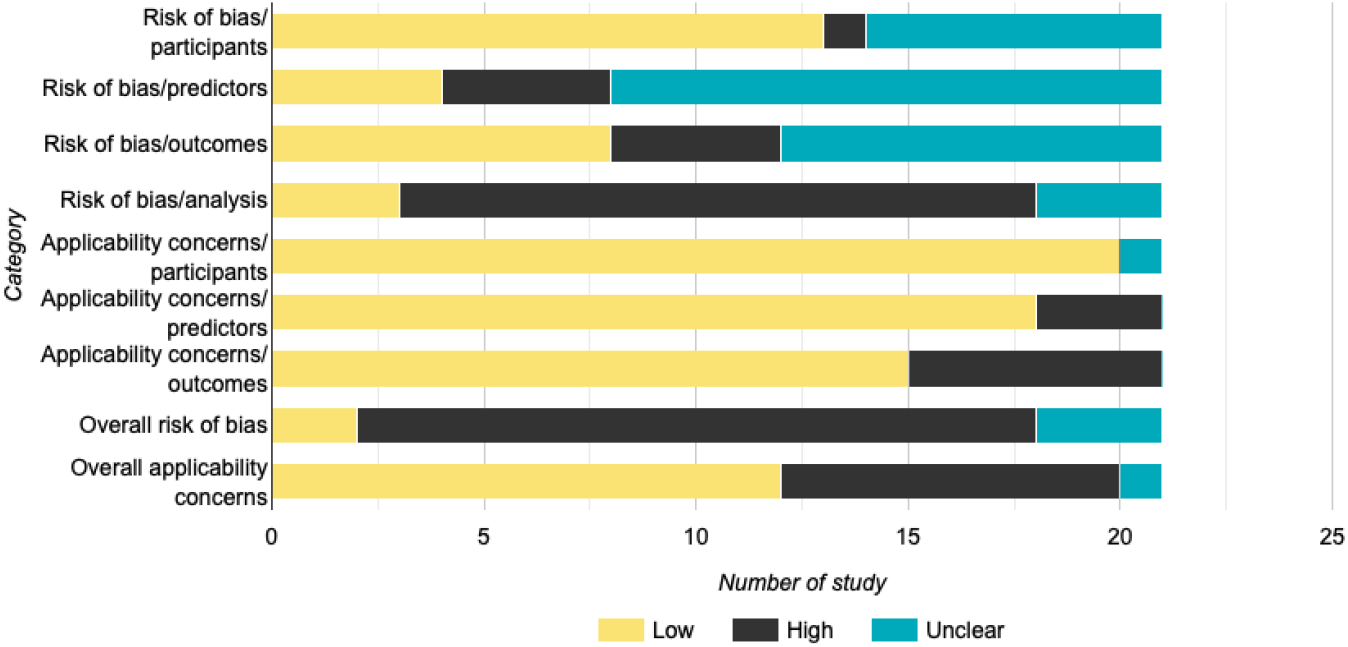
Risk of Bias and Concern or Applicability Restricted to Multi-sites

Two studies (4.3%) reported the prediction from images and seven (15%) studies reported the combinations of clinical data and images. These studies included chest X-ray and Computed Tomography (CT findings) either by itself or in conjunction with clinical data. The predictive ability of these models (AUC) reached 0.8 to 0.9, but most of these studies had a high or unclear risk of bias, which was similar to the recent review reported by Wynants et al.^11^

Overall, we identified two studies with a low risk of bias and low concern for applicability based on the PROBAST tool, which were Clift et al. and Knight et al. (Table 3)^13,15^

### Individual Study Characteristics

Detailed characteristics of each of the 46 studies are presented in Table 1. Importantly, 5 (11%) of studies used external validation of the existing clinical prediction scores, and 41 (89%) of studies developed a new model and internally or externally validated it. Two studies (4.3%) reported the prediction from images and 7 (15%) reported the combinations of clinical data and images.

### Synthesis of Results

Based on the PROBAST tools, we listed the studies with low risk of bias, low concerns for applicability, and predictive ability, metrics, setting, geographic location, and the link to an online calculator in Table 2.

**Table 2.**
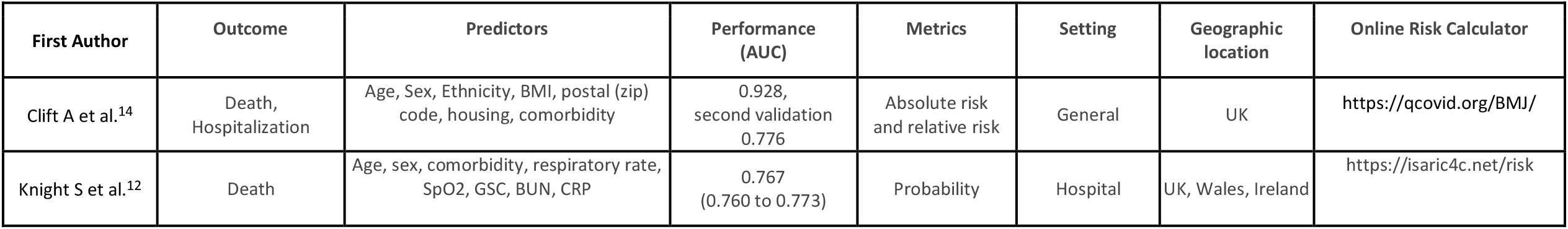
A list of studies with low risk of bias and low concern for applicability.

Two studies were found to have low risk of bias and low concern for applicability (Table 2), and therefore warrant further description. Clift et al.^15^ developed a prediction model using various risk factors including age, sex, ethnicity, BMI, postal (zip) code, housing, and comorbidity. It was conducted in the general population in the UK during the first wave, and the absolute risk depends on the incidence in the UK at the time. An example shows a 55-year-old black African man with type 2 diabetes, a body mass index of 27.7, and no other risk factors. His absolute risk of catching and dying from covid-19 over the 90 day period is 0.1095% (or 1 in 913). His relative risk compared with a white man aged 55 years and a body mass index of 25 is 10.84.

The model shows that he is in the top 10% of the population at the highest risk. Knight et al.^13^ developed and validated a risk score based on common parameters that are available at hospital admission. This included factors such as age, sex, comorbidity, respiratory rate, SpO2, GSC, BUN, and CRP. The study was on the inpatient population, and study findings, particularly the probability of mortality, could easily be adapted for use outside the UK. The study reported the low, intermediate, high, and very high risk for mortality based on predictors. For example, patients in the intermediate-risk group (score 4-8, 21.9%) had a mortality rate of 9.9%. Patients in the high-risk group (score 9-14, n=11 664, 52.2%) had a mortality rate of 31.4%.^13^ However, one should account for differences in the criteria for admission and the quality of care (Table 2).

## DISCUSSION

This systematic review identified a total of 46 articles that predicted severe COVID-19 in the literature. The majority of articles are from China, Europe, and North America where COVID-19 hit the hardest. These prediction models are focused on prognosticating patients with COVID-19. Overall, models reported the validation AUC from 0.54 to 0.98, but risk of bias was unclear or high for many studies, likely due to limited transparency on the predictor and outcome assessment, limited number of samples and outcomes. Many of these studies are retrospective studies and from early in the pandemic; most of them are limited due to the event rate and unclear case and outcomes definitions. Sample size was variable throughout studies, and lastly, analysis domain, such as overfitting were often seen as the limitation. These are commonly identified problems with building prediction models, and likely causes overfitting.^16^

Although many of the studies are reported out of urgency, several of them did not seem to follow TRIPOD guidelines,^17^ which likely contributed to the overall risk of bias for the included studies. After review of the current literature, our impression is that many of the reported prediction models have high/unclear risk of bias, thus it is not recommended that any particular tool should be used until further external validation is completed, unless the risk of bias is low.

This review identified several implications for potential application of these prediction models in the appropriate clinical settings. As stated earlier, there are 41 studies that reported original models during the pandemic, and four studies ^24,32,41,45^ that reported the external validation models from the severity prediction models which existed before the COVID-19. Clift et al., Knight et al., reported the models that were developed from nationwide data which enabled robust re-sampling methods to minimize overfitting.^13,15^ We conclude that two of them^13,15^ can be useful tools (Table 3).

Clift et al^15^ reported population-based risk algorithm in the UK, showing high levels of discrimination for deaths and hospital admissions due to covid-19. The absolute risks presented, however, will change over time in line with the prevailing SARS-CoV-2 infection rate and the extent of social distancing measures in place, so they should be interpreted with caution. Because the incidence of COVID19 varies widely according to region and timing and therefore generalizability of the absolute risk is limited. The authors reported relative risk, which is less sensitive to incidence, so that it can be used in other countries. Knight et al.^13^ reported mortality estimates from hospitalized adults in the UK. The authors categorized the risk group into very low, low, intermediate, high, and very high for in-hospital mortality. This study took place when the first wave hit the UK, and to account for concerns regarding the level of care at the time of the pandemic, the authors employed temporal validation. The prediction model could be used to determine the level of care, such as discharge or hospitalization and escalation of care after hospitalization.

### Strength and Limitations

Our review used a clear inclusion and exclusion criteria, which likely led to almost perfect agreement between two reviewers. Furthermore, the use of PROBAST strengthened our review, as this tool enabled accurate evaluation of risk of bias and applicability for each of the included studies.

There are several limitations. First, we used only two databases to identify the literature, which is less than we need for a traditional systematic review. We undertook this review to disseminate a rapidly progressing science of COVID-19 for healthcare providers. Second, our search was completed in July 2020, but this review required additional time to train reviewers to use the PROBAST tool to evaluate each article. Due to the rapid development of COVID-19 pandemic, it is important to note the gap between our search timeline and the current status at the time of article publication. Third, variables and types of prediction models were highly variable among studies, and it was infeasible to conduct meta-analysis or meta-regression, as we specified in the protocol. Our aim was to systematically curate the current state of scientific knowledge to predict severe COVID-19, and we argue that our findings are still informative to healthcare providers without a pooling of results. Fourth, we were primarily interested in multivariable models, which may not have captured innovative studies focused on one predictive marker. Lastly, the volume of literature related to this topic area was a challenge, particularly for preprint articles. Due to concerns with feasibility, database search for preprints were limited to the records included in the mentioned databases.

## CONCLUSION

Our review identified several prognostic models for COVID-19, and they showed various discriminative performances. The risk of bias was overall high or unclear for most of the studies, and the risk of bias was highest for the analysis domain. Concern for applicability was low for the majority of included studies. Thus, it is possible that most studies are prone to bias and require external validation before dissemination. The use of prediction models for severe COVID-19 cases requires caution since risk of bias is not negligible. We reported two studies that had a low risk of bias and low concern for applicability, one from a general public population and hospital setting in the UK.

## Data Availability

Per request.

## Acknowledgements

We thank the assistance to generate search strategy and identifying the literature from Jennifer Deberg, MLS, and Hans R House, MD, at the University of Iowa.

---

Supplemental file 1. Search terms for PubMed.

\# 1.

Risk Factors”[Mesh] OR “Prognosis”[Mesh] OR “ROC Curve”[Mesh] OR “Severity of Illness Index”[Mesh] OR “Hospitalization”[Mesh] OR “Forecasting”[Mesh] OR “Intubation, Intratracheal”[Mesh] OR “Respiration, Artificial”[Mesh] OR “Multiple Organ Failure”[Mesh] OR “Heart Failure”[Mesh] OR “Renal Insufficiency”[Mesh] OR “Diabetic Ketoacidosis”[Mesh] OR “Brain Diseases”[Mesh] OR “Endocrine System Diseases”[Mesh] OR predict*[Title] OR clinical score[Title/Abstract] OR risk score[Title/Abstract] OR disease severity[Title/Abstract] OR mortality[Title] OR mechanical ventilation[Title/Abstract] OR organ failure[Title/Abstract] OR heart failure[Title/Abstract] OR renal failure[Title/Abstract] OR endocrinopathy[Title/Abstract] OR encephalopathy[Title/Abstract] OR intubation[Title/Abstract]

\# 2.

covid[Title/Abstract] OR covid19 [TItle/Abstract] OR coronavirus2 [Title/Abstract] OR corona virus 2 [Title/Abstract] OR coronavirus 2 [Title/Abstract] OR corona virus 2 [Title/Abstract] OR SARS COV2 [Title/Abstract] OR SARS COV 2[Title/Abstract] OR 2019 nCOV infection[Title/Abstract] OR HCOV [Title/Abstract] OR Wuhan seafood market pneumonia virus[Title/Abstract] OR wuhan coronavirus [Title/Abstract] OR “COVID-19” [Supplementary Concept] OR “severe acute respiratory syndrome coronavirus 2” [Supplementary Concept] OR “Coronavirus Infections”[Mesh:NoExp] OR “Coronavirus”[Mesh:NoExp]

\# 3.

“Animals”[Mesh] NOT (“Animals”[Mesh] AND “Humans”[Mesh])

\# 4.

“Letter” [Publication Type] OR “Editorial” [Publication Type] OR “Comment” [Publication Type] OR “Review” [Publication Type]

#1 AND #2 NOT (#3 OR #4)= 2896, limited to english[Filter] AND 2020:2020[pdat]

